# Mitigating the impact of COVID-19 on tuberculosis and HIV services: a cross-sectional survey of 669 health professionals in 64 low and middle-income countries

**DOI:** 10.1101/2020.10.08.20207969

**Authors:** Mishal S. Khan, Sonia Rego, Joaquín Benítez Rajal, Virginia Bond, Razia Kaneez Fatima, Afshan Khurshid Isani, Jayne Sutherland, Katharina Kranzer

## Abstract

**Objective:** The experiences of frontline healthcare professionals are essential in identifying strategies to mitigate the disruption to healthcare services caused by the COVID-19 pandemic.

**Methods:** We conducted a cross-sectional study of TB and HIV professionals in low and middle-income countries (LMIC). Between May 12 and August 6 2020, we collected qualitative and quantitative data using an online survey in 11 languages. We used descriptive statistics and thematic analysis to analyse responses.

**Findings:** 669 respondents from 64 countries completed the survey. Over 40% stated that it was either impossible or much harder for TB and HIV patients to reach healthcare facilities since COVID-19. The most common barriers reported to affect patients were: fear of getting infected with SARS-CoV-2, transport disruptions and movement restrictions. 37% and 28% of responses about TB and HIV stated that healthcare provider access to facilities was also severely impacted. Strategies to address reduced transport needs and costs – including proactive coordination between the health and transport sector and cards that facilitate lower cost or easier travel - were presented in qualitative responses. Access to non-medical support for patients, such as food supplementation or counselling, was severely disrupted according to 36% and 31% of HIV and TB respondents respectively; qualitative data suggested that the need for such services was exacerbated.

**Conclusion:** Patients and healthcare providers across numerous LMIC faced substantial challenges in accessing healthcare facilities, and non-medical support for patients was particularly impacted. Synthesising recommendations of frontline professionals should be prioritised for informing policymakers and healthcare service delivery organisations.

## Introduction

The direct health effects of the COVID-19 pandemic are colossal, and are continuing to grow, with approximately one million deaths directly attributed to COVID-19 (1). However, researchers and practitioners have already highlighted that the indirect effects of COVID-19 on global health - through the disruption of essential healthcare services - may be even larger and longer lasting (2, 3).

TB and HIV are the two infectious diseases that cause the highest number of deaths globally; in 2018, 1.5 and 1.1 million people died from TB and from HIV-related illnesses respectively (5, 6). National programmes for controlling these diseases already face immense challenges, and the pandemic has increased these by diverting healthcare professionals and resources to contain COVID-19 (7, 8). Evidence from the Ebola crisis provides a warning of the reversals in progress that accompany a pandemic; for example, significant decreases in diagnoses of smear-positive TB, HIV testing and antiretroviral therapy uptake in Liberia have been documented (9). A modelling study estimating the impact of severe disruptions to service delivery predicted that HIV and TB deaths could increase by up to 10% and 20% over five years respectively in high-burden settings, reverting to levels seen a decade ago(10).

Policies to minimise disruptions to TB and HIV care must be put in place urgently. To do this, we need to understand the range of impacts of COVID-19 on TB and HIV services, and identify feasible strategies for mitigation. Information from frontline health professionals and researchers is invaluable and often insufficiently incorporated into policy planning or in the formation of research questions (11, 12). In light of this, our study rapidly synthesised information from professionals working in affected countries, in order to identify both how COVID-19 is impacting TB and HIV services in LMIC and their recommendations for minimising disruptions.

## Methods

We analysed quantitative and qualitative data collected through a rapid cross-sectional survey of TB and HIV healthcare delivery, management and research professionals in LMIC around the world.

### Study design, population and sampling

Our open online survey was conducted between May 12 and August 6 2020. The methodology was designed based on a standardised checklist for internet surveys (CHERRIES) (13). Our target population was individuals who were involved in managing or delivering TB or HIV services, including, but not limited to: doctors, nurses, community healthcare providers, laboratory technicians, policymakers, health facility managers, representatives of charity, community or advocacy groups, and researchers.

We used three approaches to share the invitation to complete our survey. First, we sent information through online professional platforms and personal networks with colleagues (using email, WhatsApp or Twitter). Second, we hired one focal point for Asia, one for Latin America, and one for Africa. Focal points focused on contacting local organisations in their regions. Third, we used snowball sampling, whereby survey participants were asked to share the survey with others who might have information to contribute. Through these approaches, the survey was shared with over 250 professional networks and organisations.

### Survey

The survey was initially designed in English by an international team with diverse expertise in TB and HIV control working in Europe, Asia and Africa. It was first piloted with eight professionals working in Cambodia, The Gambia, South Africa, The Philippines, Pakistan, Zambia and Zimbabwe; this allowed us to check face validity and refine the wording of questions and response options. Following this, we piloted an electronic version (in SurveyMonkey) with five public health professionals to check the accessibility and functionality of the survey as it appeared on the online platform.

Using SurveyMonkey enabled us to administer a survey that could only be answered once per device, and did not collect identifying information. We provided introductory text and a downloadable information sheet on the landing page. Respondents gave consent - after reading the information sheet - by checking boxes to confirm that they agreed to participate and allowed text responses to be quoted verbatim. It was possible for participants to agree to participate, but to decline use of verbatim quotes.

We used adaptive questioning, whereby nine HIV or TB-specific questions, or 18 covering both, were displayed based depending on which areas the respondent wanted to provide information about. Of the nine questions, seven were multiple choice (Table 2), with space to add free text using the ‘other’ option. There were two open response questions that only collected free text and these constitute the qualitative responses. The first asked about COVID-19 related rules that have been introduced by the government and how these have impacted TB/HIV health services. The second solicited information about measures that can be taken (or have already been taken) to minimize disruptions. There was no limit to the amount of text a respondent could insert.

**Table 1:**
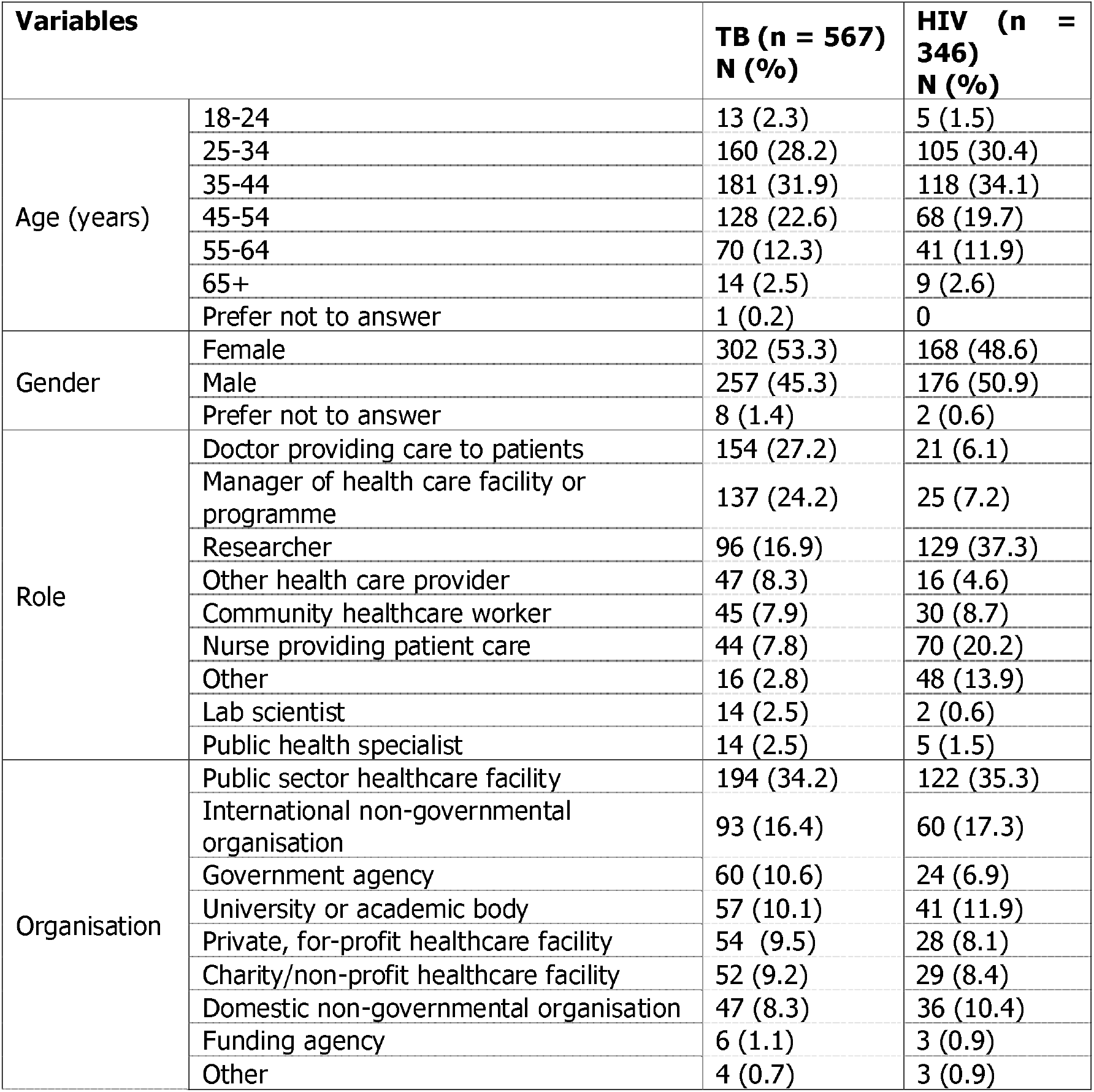
Respondents’ characteristics.

| Variables |  | TB (n = 567)<br>N (%) | HIV (n = 346)<br>N (%) |
| --- | --- | --- | --- |
| Age (years) | 18-24 | 13 (2.3) | 5 (1.5) |
|  | 25-34 | 160 (28.2) | 105 (30.4) |
|  | 35-44 | 181 (31.9) | 118 (34.1) |
|  | 45-54 | 128 (22.6) | 68 (19.7) |
|  | 55-64 | 70 (12.3) | 41 (11.9) |
|  | 65+ | 14 (2.5) | 9 (2.6) |
|  | Prefer not to answer | 1 (0.2) | 0 |
| Gender | Female | 302 (53.3) | 168 (48.6) |
|  | Male | 257 (45.3) | 176 (50.9) |
|  | Prefer not to answer | 8 (1.4) | 2 (0.6) |
| Role | Doctor providing care to patients | 154 (27.2) | 21 (6.1) |
|  | Manager of health care facility or programme | 137 (24.2) | 25 (7.2) |
|  | Researcher | 96 (16.9) | 129 (37.3) |
|  | Other health care provider | 47 (8.3) | 16 (4.6) |
|  | Community healthcare worker | 45 (7.9) | 30 (8.7) |
|  | Nurse providing patient care | 44 (7.8) | 70 (20.2) |
|  | Other | 16 (2.8) | 48 (13.9) |
|  | Lab scientist | 14 (2.5) | 2 (0.6) |
|  | Public health specialist | 14 (2.5) | 5 (1.5) |
| Organisation | Public sector healthcare facility | 194 (34.2) | 122 (35.3) |
|  | International non-governmental organisation | 93 (16.4) | 60 (17.3) |
|  | Government agency | 60 (10.6) | 24 (6.9) |
|  | University or academic body | 57 (10.1) | 41 (11.9) |
|  | Private, for-profit healthcare facility | 54 (9.5) | 28 (8.1) |
|  | Charity/non-profit healthcare facility | 52 (9.2) | 29 (8.4) |
|  | Domestic non-governmental organisation | 47 (8.3) | 36 (10.4) |
|  | Funding agency | 6 (1.1) | 3 (0.9) |
|  | Other | 4 (0.7) | 3 (0.9) |

**Table 2:** Quantitative survey findings.

| **Questions** |  | **TB (n = 567)**<br>**N (%)** | **HIV (n = 346)**<br>**N (%)** |
| --- | --- | --- | --- |
| Has it been harder for healthcare providers to come to work at healthcare facilities since COVID-19? | No- same as before | 98 (17.3) | 61 (17.6) |
|  | Yes - it is slightly harder | 245 (43.2) | 182 (52.6) |
|  | Yes - it is much harder | 179 (31.6) | 85 (24.6) |
|  | Yes - it is very difficult or impossible | 28 (4.9) | 13 (3.8) |
|  | Don't know | 14 (2.5) | 4 (1.2) |
|  | Prefer not to answer | 3 (0.5) | 1 (0.3) |
| Has it been harder for patients to come seek care at healthcare facilities since COVID-19? | No- same as before | 56 (9.9) | 37 (10.7) |
|  | Yes - it is slightly harder | 266 (46.9) | 159 (46.0) |
|  | Yes - it is much harder | 193 (34.0) | 120 (34.7) |
|  | Yes – it is very difficult or impossible | 40 (7.1) | 24 (6.9) |
|  | Don't know | 8 (1.4) | 5 (1.5) |
|  | Prefer not to answer | 4 (0.7) | 1 (0.3) |
| What do you think are the main concerns or barriers for patients to access healthcare since COVID-19 | Fear of getting infected with SARS-CoV-2 | 422 (74.4) | 247 (71.4) |
|  | Transport disruption | 395 (69.7) | 270 (78.0) |
|  | Lockdown | 388 (68.4) | 207 (59.8) |
|  | Reduced income/ money to travel | 295 (52.0) | 260 (75.1) |
|  | Clinical closures | 142 (25.0) | 97 (28.0) |
|  | Health care workers shortage | 131 (23.1) | 102 (29.5) |
|  | Unable to access a face mask | 123 (21.7) | 68 (19.7) |
|  | Long waiting times | 91 (16.0) | 80 (23.1) |
|  | There are no concerns | 48 (8.5) | 29 (8.4) |
| Since COVID-19, are you aware of any changes to the way healthcare facilities are operating? | Physical distancing protocols for patients | 420 (74.1) | 272 (78.6) |
|  | Masks or other protective equipment for healthcare providers | 399 (70.4) | 255 (73.7) |
|  | Prefer not to answer | 22 (3.9) | 12 (3.5) |
| Have you experienced shortages of diagnostics or other challenges to provision of routine diagnostic services since COVID-19? | No- same as before | 168 (29.6) | 120 (34.7) |
|  | Yes - it is slightly harder | 193 (34.0) | 124 (35.8) |
|  | Yes - it is much harder | 139 (24.5) | 63 (18.2) |
|  | Yes – it is very difficult or impossible | 23 (4.1) | 19 (5.5) |
|  | Don't know | 36 (6.3) | 16 (4.6) |
|  | Prefer not to answer | 8 (1.4) | 4 (1.2) |
| Have you experienced shortages of medicines or other challenges to provision of standard treatment since COVID-19? | No- same as before | 300 (52.9) | 159 (46.0) |
|  | Yes - it is slightly harder | 150 (26.5) | 113 (32.7) |
|  | Yes - it is much harder | 58 (10.2) | 43 (12.4) |
|  | Yes – it is very difficult or impossible | 7 (1.2) | 13 (3.8) |
|  | Don't know | 41 (7.2) | 16 (4.6) |
|  | Prefer not to answer | 11 (1.9) | 2 (0.6) |
| Has it been harder for patients to access non-medical support such as food supplementation or counselling since COVID-19? | No- same as before | 103 (18.2) | 60 (17.3) |
|  | Yes - it is slightly harder | 223 (39.3) | 122 (35.5) |
|  | Yes - it is much harder | 137 (24.1) | 97 (28.0) |
|  | Yes – it is very difficult or impossible | 38 (6.7) | 27 (7.8) |
|  | Don't know | 19 (3.4) | 6 (1.7) |
|  | Prefer not to answer | 47 (8.3) | 27 (7.8) |

The survey and participant information sheet were translated into 10 additional languages: Arabic, Bahasa, Chinese, French, Portuguese, Russian, Shona, Spanish, Swahili, and Urdu. There were at least two translators for each language, so that every translation was checked by an independent native or fluent speaker.

### Data management and analysis

All data was downloaded into MS Excel. The quantitative data was analysed using descriptive statistics (frequencies and percentages) in Stata/SE V.14 (StatCorp, Texas, USA).

Our qualitative analysis involved translating non-English language text into English, and conducting a thematic analysis. We used an interpretive approach in which identified themes were supported by the data. The thematic analysis process began by two authors agreeing on the emerging themes after reading the text responses independently and then coding the text line by line manually. Finally, we triangulated qualitative and quantitative results to validate findings.

### Ethical Approval

We received ethical approval from the London School of Hygiene and Tropical Medicine, the University of Zambia and The South African Medical Association.

## Results

Of the 923 respondents that initiated the consent process and accessed survey questions, 669 (72%) from 64 countries completed it. 567 respondents answered the TB section, and 346 answered the HIV section. There was a greater representation of health professional based in sub-Saharan African countries in responses about HIV as compared to TB (Figure 1). Demographic and professional characteristics of respondents are in Table 1.

**Figure 1:**
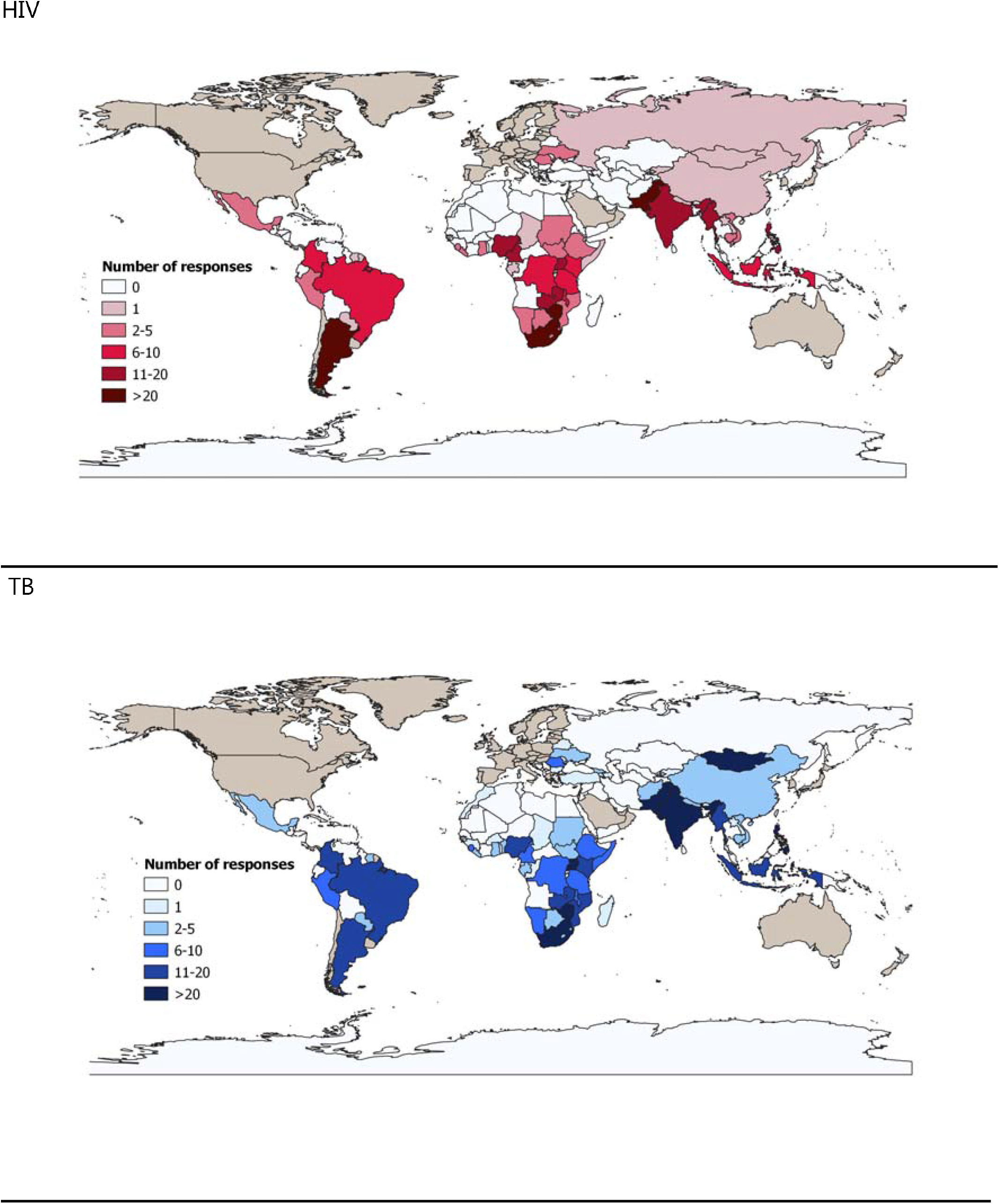
Geographical distribution of survey respondents.

### Access to healthcare facilities for patients and providers

Our survey indicated that access to healthcare facilities for TB and HIV healthcare providers and patients has been substantially affected (Table 2). Over 40% of respondents stated that it was impossible or much harder for TB and HIV patients to reach healthcare facilities since COVID-19. Similarly, it was much harder or impossible for TB healthcare providers to reach their place of work since COVID-19 began, according to 37% of respondents. Challenges were also reported in relation to HIV healthcare providers reaching their place of work, but these were not considered as severe as for HIV patients.

Concerns and barriers for TB and HIV patients to access healthcare were similar. The following were identified by more than 50% of respondents: fear of getting infected with SARS-CoV-2, transport disruptions, movement restrictions owing to lockdowns and reduced income. Disruption to transport services was a recurring theme in the qualitative data, mentioned over 30 times across Asian, African and Latin American countries. Specifically, it was highlighted that both reduced access to usual modes of transport and increased cost of transport created barriers to reaching healthcare facilities for both healthcare providers and patients. Furthermore, the impact of transport disruptions on essential supplies reaching healthcare facilities was noted repeatedly as an issue to address: *Monitor the lockdown enforcement to make sure it really doesn’t hamper movement of essential items; other endemic diseases should not be neglected due to COVID-19. [M, doctor, Nigeria]*. Three policies to address transport disruptions suggested were: proactive coordination between the health and transport sector (including private transport companies); cards that facilitate lower cost or easier travel for patients and providers, and public information to reassure patients that they can travel for healthcare.

Alternative solutions, suggested by over 40 respondents, focused on ways to reduce the need for patients to travel to healthcare facilities. These solutions relied on bringing services closer to patients’ homes, and providing medication for longer durations. As an adaptation to COVID-19 situation, it was commonly reported that medicines for TB and HIV were allowed to be given for several months at one time; some responses indicated challenges in doing this due to insufficient supplies. Provision of HIV and TB medicines to cover longer periods, however, does not address the need for patients to interact with healthcare providers as part of treatment monitoring and follow-up support. Telemedicine was identified as a solution by more than 50 respondents. This was reported to be operating in some facilities in countries such as Argentina and South Africa, and was proposed as a way of providing better services in the future, even outside of a health emergency. *Antiretrovirals are handed out at home to avoid patient exposure*…*a telephone line was implemented for access to consultations through telemedicine*. [F, manager of health facility, Argentina]

Recommendations to bring healthcare closer to communities included delivery of counselling and medicines through community volunteers or local private providers, and setting up community collection points. Numerous respondents encouraged flexibility in where patients are allowed to collect medicines, since some patients may relocate to their villages from urban areas when employment opportunities are affected.

### Disruptions to service provision at TB and HIV facilities

Over 70% of respondents reported that TB and HIV facilities had revised their operating procedures to include physical distancing protocols for patients, and over 70% also reported that TB and HIV facilities had introduced Personal Protective Equipment (PPE) for healthcare providers. The importance of PPE, and masks in particular, was mentioned commonly (35 times for healthcare providers and 20 times for patients) across numerous countries. We noted a difference in whether respondents stressed the need for masks specifically for doctors, or for other providers such as laboratory technicians and community healthcare workers. We also found that some doctors and healthcare managers emphasised the need for education on infection prevention practices in conjunction with increased access to masks. Concerns were raised about a lack of clarity on who should be responsible for providing PPE, and the cost burden on poorer patients if they are required to source their own masks: *Most TB patients are poor. They cannot afford masks and sanitizer [F, researcher, Pakistan]*

Only 16% and 11% of HIV and TB respondents reported that standard medical treatment was very difficult or impossible to provide. In contrast, we found that access to non-medical support for patients, such as food supplementation (where available) or counselling, was much harder or impossible to access according to 36% and 31% of respondents that answered questions about HIV and TB respectively. Less than 20% said that these important services were unchanged. Furthermore, the qualitative data suggested that nutritional support became particularly critical when movement restrictions and employment instability further reduced access to income and food. A repeated suggestion was that food supplementation should be paired with community-based delivery of drug supplies: *Poor patients also miss the nutrition they used to receive at the TB facilities. Provide nutrition with the drug supplies [M, health facility manager, Somalia]*

While many respondents advocated for the upkeep of counselling services for patients, the need for mental health support for providers was not mentioned. Increased workload and stress was mentioned as a problem, but few provided solutions. The solutions mentioned included financial incentives to compensate for the increased occupational risk and hiring of additional healthcare providers to account for the greater work load. Shift working systems were also suggested to reduce the number of healthcare providers in facilities at any given time, especially in space-limited settings such as laboratories.

### Impacts on supply chains and diagnostic services

There were mixed views on challenges around the provision of routine diagnostic services for TB and HIV, indicating that the experiences of respondents varied considerably. Approximately one third reported that providing diagnostic services had been slightly harder and a similar proportion reported no change since COVID-19, while 29% and 24% reported that TB and HIV diagnostic services were very hard or impossible to provide. Qualitative data indicated that countries with more centralised production or storage of essential supplies might face greater challenges in maintaining supply chains for diagnostics due to greater reliance on transport. Similarly, where diagnostics were not produced domestically, risk of disruptions increased, according to the qualitative data. Suggested solutions included shifting to quality-assured local production and decentralisation of diagnostics wherever possible.

### Stigma

Qualitative responses highlighted increased stigmatisation of HIV and TB patients owing to changes in the delivery of health services. Examples provided by respondents included stigmatisation when HIV patients were asked to show health cards in order to travel, when patients presenting with TB symptoms were first isolated and tested for COVID-19, and when attention is drawn to HIV or TB patients in their neighbourhoods during community-based distribution of medicines.

*(It is) difficult to ask people living with HIV to show their medical records at road blocks as this discloses their status. [F, doctor, Zimbabwe]*

*To develop a strategy for the supply of medication at a community level, being supplied together with other types of assistance to protect the identity of those wishing for health status anonymity. [F, community healthcare worker, Dominican Republic]*

Another strong theme was fear and stigma of COVID-19. It was frequently reported that people were worried that healthcare providers or community members would think they had COVID-19 if they sought care for TB. This was identified as a priority area to be addressed: *Need to reduce stigma about COVID-19 and associations with TB. [M, manager, Kenya]*

## Discussion

Our multi-country survey provides current evidence of the widespread impacts of COVID-19 on TB and HIV patients, healthcare providers and delivery of routine services. It also identifies specific policies and service delivery adaptations that can be implemented to mitigate disruptions. We summarise three key disruptions and their implications. First, we found that disruptions to transport services posed substantial challenges not only to patients, but also to healthcare providers. Second, our data indicated that addressing transport challenges alone would not be sufficient, owing to the barrier posed by patients’ fears of contracting SARS-CoV-2. Third, in terms of disruption to service provision, the reduction in patient access to critical non-medical support was striking. Less than 20% of respondents stated that access to non-medical support was unaffected. As suggested by our qualitative data and other studies (14-16), mitigating disruptions to provision of food supplementation and mental health support is critical due to the increased stress, unemployment and loss of income caused by health emergencies.

Frontline professionals identified important solutions to minimise disruptions from health emergencies. Health service adaptations to reduce the need for long journeys to healthcare facilities, included: community-based kiosks to enable collection of medication, telemedicine services and telephone helplines. Some of these were proposed as sustainable improvements for healthcare delivery that could be adopted even outside of emergency situations. Practical strategies to address barriers to using transport services for accessing healthcare and to improve healthcare provider motivation and safety were also identified.

Researchers have concluded that health system resilience entails a combination of absorptive, adaptive and transformative strategies (20). For example, overcrowding of urban health can at least partly be overcome by community-based care and telemedicine. However, the consequences of ongoing adaptations on health outcomes and the impacts of changes in service delivery on stigma and equity must be constantly examined. For example, home delivery of medications may impact the former, while reliance on access to a phone for telemedicine services may impact the latter.

Complex health systems consist of both hardware(infrastructure, commodities, human resources and finances) and software (knowledge, values, norms, and feelings that shape health service delivery) (21). Our study indicated that the software of health systems, such as mental health and motivation of healthcare workers, both of which are known to be affected in outbreak situations, was being neglected by current responses to COVID-19 (22). Finally, our study demonstrates that frontline professional in LMICs have invaluable experience, and that sharing information between organisations in the same country and across LMIC should be facilitated. Solutions applied in one country could be adapted for use in another LMIC, and may be more appropriate than policies designed by decision makers who are less embedded in the realities of LMIC health services (17-19).

Although there are important strengths of our survey-based study, such as rapid gathering of data in low technology settings, and the wide array of countries we received data from, we acknowledge that bias can result from 1) the non-representative nature of the population sampled and 2) the self-selection of participants. Though the survey was available in 11 languages, we missed professionals who speak other languages and we were informed by colleagues in some countries (Russia and China) that participants did not feel comfortable responding to such a survey. We also received more responses about TB than HIV.

## Conclusions

Data from this first multi-country survey focusing on experiences of frontline professionals showed that challenges to accessing healthcare facilities and maintaining routine service delivery – particularly in relation to diagnostics and non-medical support - were substantial across LMIC following COVID-19. Frontline professionals identified important mitigation strategies, including adaptations to reduce the costs or need for patients and providers to travel to healthcare facilities, measures to address healthcare provider safety and motivation, and approaches to tackle increases in stigma. These professionals, who are deeply involved in delivering, managing or analysing service delivery during emergencies such as COVID-19, will not have the time or connections to influence planning of policies, despite having insights that are critical for effective policy setting. Rapid synthesis of information them can facilitate identification of service delivery barriers and bottlenecks presenting during emergency situations, and help develop effective adaptations. Furthermore, some of the emerging changes to healthcare delivery models could support better resilience against future emergencies.

## Data Availability

Data about responses from individual countries cannot be shared publicly because small numbers of responses from certain countries lead to the risk of respondents being identifiable. We are committed to sharing data in line with the rules of our ethical approval. Data are available from the corresponding author for researchers who meet the criteria for access to confidential data.

## Acknowledgements

We had over twenty volunteers help us pilot and translate the survey and we are grateful to each of them. We thank Chepela Ngulube for supporting us on the ethics submission in Zambia. We would also like to acknowledge the LSHTM Alumni Association and LSHTM TB Centre.

